# Mask-associated de novo headache in healthcare workers during the Covid-19 pandemic

**DOI:** 10.1101/2020.08.07.20167957

**Authors:** José M Ramírez-Moreno, David Ceberino, Alberto González-Plata, Belen Rebollo, Pablo Macías-Sedas, Roshu Hariranami Ramchandani, Ana M Roa, Ana B Constantino

## Abstract

**Introduction:** The pandemic caused by the new coronavirus (COVID-19) has led to changes in the development of health care activities by health professionals. We analysed whether there is an association between the appearance of “de novo” headache according to the type of mask used, the related factors, as well as the impact of the headache on health professionals.

**Method:** cross-sectional study in a tertiary hospital in Extremadura, Spain. We administered an online questionnaire to healthcare workers during the period of maximum incidence of COVID-19 in our setting.

**Results:** n=306, 244 women (79.7%), with an average age of 43 years (range 23-65). Of the total, 129 (42.2%) were physicians, 112 (36.6%) nurses and 65 (21.2%) other health workers. 208 (79.7%) used surgical masks and 53 (20.3%) used filtering masks. Of all those surveyed, 158 (51.6%) presented “de novo” headache. The occurrence of headache was independently associated with the use of a filtering mask, OR 2.14 (IC95% 1.07-4.32), being a nurse OR 2.09 (IC95% 1.18-3.72) or another health worker OR 6.94 (IC95% 3.01-16.04) or having a history of asthma OR 0.29 (IC95% 0.09-0.89). Depending on the type of mask used there were differences in headache intensity. And the impact of headache in the subjects who used a filtering mask was worse in the all aspects evaluated.

**Conclusions:** The appearance of “de novo” headache is associated with the use of filtering masks and is more frequent in certain health care workers, causing a greater occupational, family, personal and social impact.

## Introduction

In December 2019, a new coronavirus, SARS-CoV-2, started an outbreak in the Chinese city of Wuhan. In January 2020 its clinical picture was defined as a disease associated with coronavirus-2019 (COVID-19) [1,2]. This outbreak has evolved into a pandemic and as of May 24, 2020, 216 countries have been affected, 5,206,614 cases have been confirmed worldwide, and 337,736 deaths have occurred [3]. In Spain, 233,037 cases have been documented and 27,940 patients have lost their lives [4]. In the region of Extremadura, 3,047 cases and 506 deaths have been reported [5].

During the increase in cases of COVID-19 in our environment, the national and local authorities established the mandatory use of Personal Protective Equipment (PPE) by health professionals. This PPE consists of a protective suit, surgical gloves, protective goggles, shield and face mask. In the case of face masks, they must be highly effective, with type FPP2 (in Europe), N95 (USA) and KN95 (China) recommended [6]. There are other types of masks (surgical masks or FPP1 among others), of lesser effectiveness, which are used by healthcare personnel who are not in direct contact with COVID-19 [7]. The use of protective material in a strict manner is crucial, as it can reduce transmission to highly exposed populations such as healthcare workers, as well as reduce the spread of infection from healthcare workers to healthy patients.

In “frontline” work, the use of masks can be very prolonged [8]. Although, in general, highly effective masks are well tolerated, some problems have been reported, such as: general discomfort, decreased visual, auditory or vocal capacity; excessive heat or humidity, facial pressure, skin lesions, itching, fatigue, anxiety and claustrophobia [9]. Another effect, already described in the 2003 SARS epidemic, was headache, whose prevalence reached 37.3% of the health personnel studied [10]. Headache associated with mask use could be related to mechanical factors, the presence of hypoxemia and hypercapnia or to the stress associated with mask use [11, 12].

Our aim is to demonstrate whether there is an association between the appearance of “de novo” headache with the type of mask and its time of use, as well as the impact of this headache on health professionals.

## Method

The study was conducted in the health area of a tertiary hospital, where our health system in the COVID period was mandated to use PPE during contact with patients.

These protective systems were mandatory among health workers, both in high-risk areas (intensive care units, isolation rooms for infected patients, emergency rooms or operating theatres), and in general medical wards, central hospital radiology and diagnostic imaging areas or outpatient clinics. This involved the use of different types of more or less tight-fitting masks, and sometimes glasses or screens.

Using a self-administered questionnaire addressed to health workers in our health area, we carried out a cross-sectional study during the first week of May 2020. In the previous month, the number of admissions for COVID-19 was very high and attendance protocols required the use of these devices by all workers.

The questionnaire collected the following information: (1) demographics (gender, age, profession, shifts); (2) medical history, including SARS-Cov2 infection; (3) type and pattern of mask use (surgical masks vs. self-filtering masks of particles and liquid aerosols (FFP), average number of hours of use per day) and use of other protective devices (glasses or screens); (4) frequency and characteristics of pre-existing primary headache (changes in headache frequency, attack duration and frequency, as well as drug use and response), (5) the main variable of the study was personal opinion about the presence of new headache in the period in which these protective systems were mandatory (duration of headache episode, intensity and frequency, as well as drug use and response); (6) presence of other symptoms potentially associated with the use of facial protection equipment (fatigue, sleep disorder, lack of concentration, irritability, nausea or vomiting or others); (7) we evaluated the self-perceived impact of the presence of new-onset headache using the Likert scale on social, occupational, family and personal aspects; (8) we also evaluated the self-perceived impact that headache conditions have on the performance of work activities and (9) lastly, we analyzed self-perceived work stress by means of the Psychosomatic Problems Questionnaire (PPQ) [13].

The questionnaire was written after an analysis of the literature and a thorough reflection on the problem to be investigated. It included a request for voluntary collaboration, information on the reason for the survey, instructions for completing the questionnaire and consent. The average time taken to complete the questionnaire was about 20 minutes.

The information collection procedure chosen was the online survey. The survey was scheduled to be conducted over five consecutive days, between 1 and 6 May 2020, with the data collected referring to the previous month.

The data collected in the study respects the anonymity of the subject and there is no possibility of access to any personal information of the individual. The data analysed is restricted to the study investigators, health authorities and the Clinical Research Ethics Committee, when required, in accordance with current legislation.

## Statistical Analysis

Prior to the analysis of relationships between variables, descriptive analyses of the different areas that make up the study have been carried out. These descriptive analyses include percentage distributions of the different categories of the analysed variables and, in the case of quantitative variables, average and standard deviation. These same analyses, shown as a cross between variables by means of contingency tables or comparison of averages, have also been elaborated as a preamble to the statistical tests that have been carried out to corroborate if there is a relationship between different variables, thus showing the hypotheses to be contrasted.

Depending on the nature of the variable (qualitative or quantitative) and the distribution of the sample (normal, admitting parametric contrasts, or non-normal, needing non-parametric contrasts), different tests have been used. We used the chi-square test to contrast whether there is independence between two categorical variables using a contingency table when the data are not paired.

For the analysis of the predictive factors with the appearance of a “de novo” headache, we used binary logistic regression methods by steps backwards, to maximize sensitivity, variables with a univariate association of P <0.200 were included as candidates in the multivariate model.

To measure the relationship between the different variables in the study, statistical tests with a 95% significance level were used as an acceptance threshold for the hypotheses to be tested, i.e. a p-value of 0.05. All statistical analyses were performed using the SPSS version 25.0 statistical package program for Windows (SPSS Inc, 2003, Chicago, IL, USA).

## Results

A total of 306 health professionals and other health workers participated in the study, 62 men (20.3%) and 244 women (79.7%), with an average age of 43 years (SD, 11; range, 23-65). Of these, 129 (42.2%) participants were physicians, 112 (36.6%) nurses and the rest, 65 (21.2%) other health workers (assistants, guards, technicians, administrative staff). With regard to the work shift, 89 (34.1%) worked in the morning and on duty, 91 (34.9%) in morning, afternoon and night shifts and 81 (31.0%) in morning shifts only. The surgical mask was used by 208 (79.7%) of those surveyed and the filtering mask (FFP2 or KN95) was used by 53 (20.3%), with no difference in the mean time of use of 7.0 (SD 2.3) hours vs 6.7 (SD 2.5) hours, p=0.289. 46.4% (121) reported not habitually using other facial protection devices such as glasses, screens or PPE. The rate of confirmed SARS-CoV 2 infection in the study population was 4.6%.

The most frequently reported diseases in the total sample in order of frequency were: allergy 34 (13.0%), thyroid diseases 28 (10.7%), anxiety 26 (10.0%), high blood pressure 18 (6.9%), asthma 17 (6.5%), dyslipemia 14 (5.4%) and diabetes 2 (0.8%). 15.7% (41) indicated tobacco consumption.

Of the 306 persons surveyed, 158 (51.6%) reported the appearance of a new headache during the period of study, of whom 65 (41.1%) had previously had a headache (migraine: 27 (17.1%), tension: 26 (16.5%) and others: 11 (6.9%)). There were 103 (33.7%) subjects who did not observe the appearance of new headache. A 14.7% were undecided on the answer (“I don’t know”) or the answer was “maybe”; these 45 subjects were eliminated from the analysis.

They were also asked about the presence of other symptoms such as sleep disturbance, loss of concentration, irritability, photophobia, sonophobia, nausea or vomiting. Table 1 shows the characteristics of the population according to the appearance or not of headache.

**Table 1.** Baseline Conditions and mask and Personal Protective Equipment usage among healthcare workers.

|  | De Novo Headache |  | P-value |
| --- | --- | --- | --- |
|  | NO<br>(n=103) | YES<br>(n=158) |  |
| Age (years) | 40.8 (11.4) | 44.4 (10.1) | 0.009 |
| Female Gender | 73 (70.9%) | 135 (85.4%) | 0.004 |
| Occupation |  |  | 0.0001 |
| Doctor | 61(59.2%) | 51 (32.3%) |  |
| Nurse | 33 (32.0%) | 62(39.2%) |  |
| Others | 9(8.7%) | 45(28.5%) |  |
| Work shift |  |  | <0.0001 |
| Mornings and 24 h. duties | 51 (49.5%) | 38 (24.1%) |  |
| Rotating shifts | 26 (25.2%) | 65 (41.1%) |  |
| Others | 26 (25.2%) | 55 (34.8%) |  |
| Type of face mask |  |  | 0.029 |
| Surgical mask | 89 (86.4%) | 119 (57.2%) |  |
| N95/FFP2 | 14 (13.6%) | 39 (24.7%) |  |
| Number of hours worn per day (SD) | 6.8 (2.4) | 7,0 (2,2) | 0.474 |
| Use of another PPE: |  |  | 0.203 |
| Face shield | 21 (20.4%) | 33 (20.9%) |  |
| Protective eyewear | 13 (12.6%) | 20 (12.7%) |  |
| Complete PPE | 16 (15.5%) | 30 (19.0%) |  |
| Confirmed COVID-19 | 4 (3.9%) | 8 (5.1%) | 0.161 |
| Pre-existing headache | 45 (43.7%) | 65 (41.1) | 0.683 |
| Comorbidity |  |  |  |
| Allergy | 16 (15.5%) | 18(11.4%) | 0.331 |
| Asthma | 11 (10.7%) | 6 (3.8%) | 0.028 |
| Tobacco | 8 (7.8%) | 33 (20.9%) | 0.004 |
| Arterial hypertension | 6 (5.8%) | 8 (5.1%) | 0.789 |
| Cardiopathy | 0 (0.0%) | 1 (0.6) | 0.419 |
| Dyslipidemia | 7 (6.8%) | 7 (4.4%) | 0.760 |
| Diabetes | 1 (1.0%) | 1 (0.6%) | 0.760 |
| Thyroid disease | 9 (8.7%) | 19 (12%) | 0.402 |
| Anxiety | 6 (5.8%) | 20 (12.7%) | 0.072 |
| Others | 4 (3.9%) | 12 (7.6%) | 0.222 |
| Other symptoms |  |  |  |
| Sleep disturbance | 11 (10.7%) | 68 (43%) | 0.0001 |
| Loss of concentration | 14 (13.6%) | 59 (37.3%) | 0.0001 |
| Irritability | 18 (17.5%) | 56 (35.4%) | 0.002 |
| Photophobia | 8 (7.8%) | 23 (14.6%) | 0.097 |
| Sonophobia | 6 (5.8%) | 18 (11.4%) | 0.128 |
| Sickness/vomiting | 11 (10.7%) | 13 (8.2%) | 0.503 |

h.: hours; PPE: Personal Protective Equipment; HCW: healthcare workers; SD: standard deviation; VAS: visual analogic scale

During April, the month immediately prior to the survey, participants with “de novo” headache presented a median of 12 (IQR 13) days of headache, median of 4 days (IQR 3) in the week prior to the survey and the pain presented an average intensity on the visual analogue scale of 6 (SD 1.5). In 74 (47.4%) subjects the duration was from 1 to 4 hours, in 46 (29.5%) from 4 to 8 hours, in 21 (13.5%) from 8 to 12 hours and in 15 (9.6%) more than 12 hours. In subjects with previous headache the duration of episodes was significantly higher (p=0.008). The response to analgesics was good or very good in 61.4% of the cases. Only 2 (1.3%) subjects had to consult the emergency department for headache, and no subject had been admitted to hospital for headache. With respect to the impact of headache in the work setting, lack of concentration on tasks was the main complaint (105 (66.5%) subjects). Table 2 shows the main characteristics of “de novo” headache.

**Table 2.**
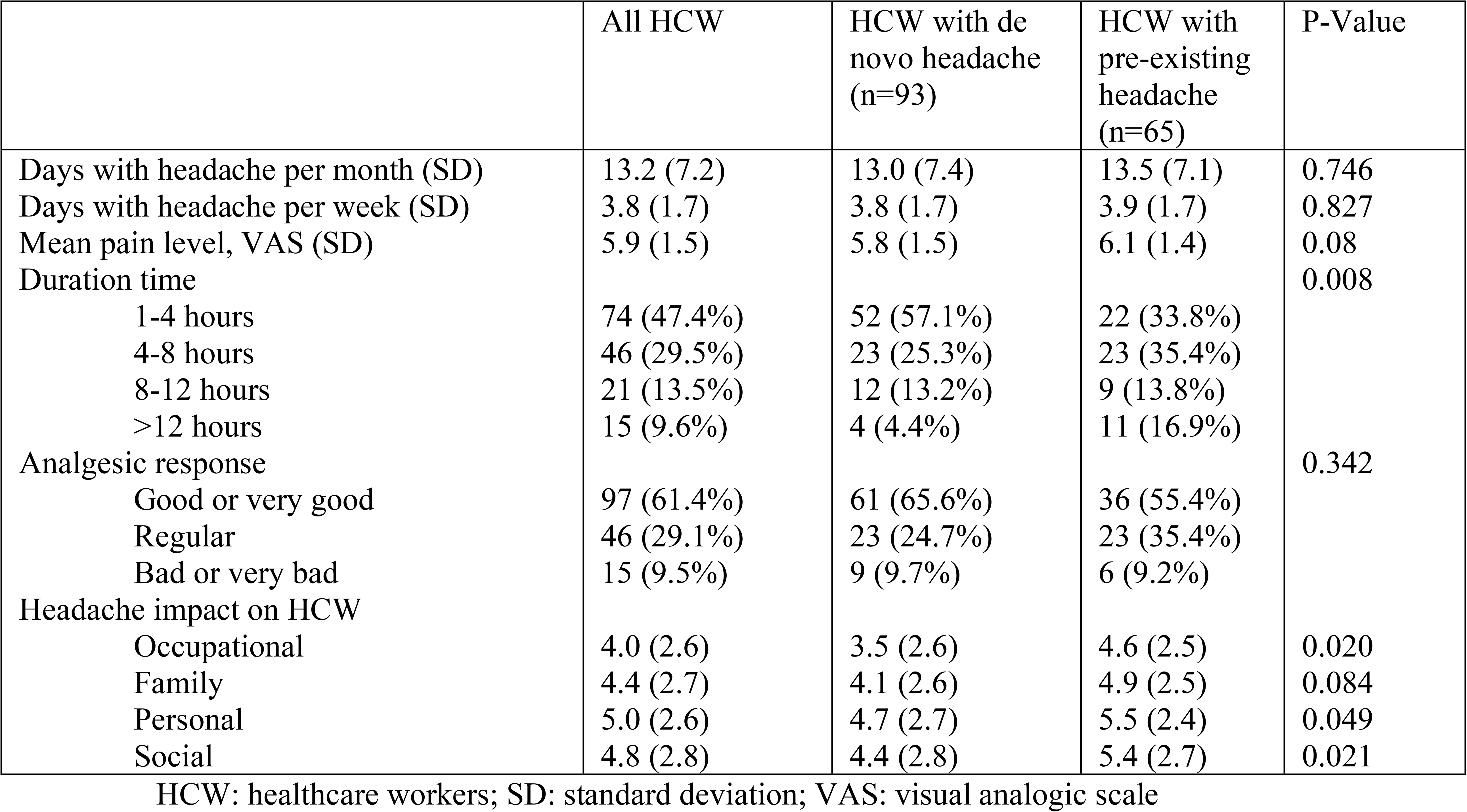
Characteristics of Headache in healthcare workers mask users.

|  | All HCW | HCW with de novo headache (n=93) | HCW with pre-existing headache (n=65) | P-Value |
| --- | --- | --- | --- | --- |
| Days with headache per month (SD) | 13.2 (7.2) | 13.0 (7.4) | 13.5 (7.1) | 0.746 |
| Days with headache per week (SD) | 3.8 (1.7) | 3.8 (1.7) | 3.9 (1.7) | 0.827 |
| Mean pain level, VAS (SD) | 5.9 (1.5) | 5.8 (1.5) | 6.1 (1.4) | 0.08 |
| Duration time |  |  |  | 0.008 |
| 1-4 hours | 74 (47.4%) | 52 (57.1%) | 22 (33.8%) |  |
| 4-8 hours | 46 (29.5%) | 23 (25.3%) | 23 (35.4%) |  |
| 8-12 hours | 21 (13.5%) | 12 (13.2%) | 9 (13.8%) |  |
| >12 hours | 15 (9.6%) | 4 (4.4%) | 11 (16.9%) |  |
| Analgesic response |  |  |  | 0.342 |
| Good or very good | 97 (61.4%) | 61 (65.6%) | 36 (55.4%) |  |
| Regular | 46 (29.1%) | 23 (24.7%) | 23 (35.4%) |  |
| Bad or very bad | 15 (9.5%) | 9 (9.7%) | 6 (9.2%) |  |
| Headache impact on HCW |  |  |  |  |
| Occupational | 4.0 (2.6) | 3.5 (2.6) | 4.6 (2.5) | 0.020 |
| Family | 4.4 (2.7) | 4.1 (2.6) | 4.9 (2.5) | 0.084 |
| Personal | 5.0 (2.6) | 4.7 (2.7) | 5.5 (2.4) | 0.049 |
| Social | 4.8 (2.8) | 4.4 (2.8) | 5.4 (2.7) | 0.021 |

HCW: healthcare workers; SD: standard deviation; VAS: visual analogic scale

83.1% (54) of the 65 subjects with previous headache indicated a modification in the characteristics of their habitual headaches, 81.0% (47) a change in location, 67.2% (39) in frequency, 36.2% (21) in intensity and 25.9% (15) in the response to habitual analgesics.

In the univariant analysis, the factors associated with the appearance of “de novo” headache were: age, female sex, type of professional, use of filter mask (KN95 or FFP2), work shift, being a tobacco user, suffering from anxiety or asthma. In the multivariant analysis, the use of filter masks and the type of professional behaved as independent predictors of headache risk, while being asthmatic behaved as a protective factor. The occurrence of headache is associated with the use of a filtering mask (FFP2 or KN95), OR 2.14 (IC95% 1.07-4.32), being a health worker OR 6.94 (IC95% 3.01-16.04) or a nurse OR 2.09 (IC95% 1.18-3.72). Table 3.

**Table 3.** Univariate and multivariate analysis of Factors of Baseline Conditions

|  | UNIVARIATE ANALYSIS |  |  |  | MULTIVARIATE ANALYSIS |  |  |  |
| --- | --- | --- | --- | --- | --- | --- | --- | --- |
| VARIABLES | OR | CI 95% |  | p-value | OR | CI 95% |  | p-value |
| Age | 1.03 | 1.01 | 1.06 | 0.009 |  |  |  |  |
| Female Gender | 2.41 | 1.31 | 4.45 | 0.005 |  |  |  |  |
| Doctor | Ref | Ref | Ref | 0.0001 | ref | ref |  | <0.0001 |
| Nurse | 2.25 | 1.28 | 3.94 |  | 2.09 | 1.18 | 3.72 |  |
| Other HCW | 5.98 | 2.67 | 13.4 |  | 6.94 | 3.01 | 16.04 |  |
| Filter mask vs Surgical | 2.08 | 1.07 | 4.07 | 0.026 | 2.14 | 1.07 | 4.32 | 0.027 |
| Mornings and 24 h duties | Ref | Ref | Ref | 0.0001 |  |  |  |  |
| Rotating shifts | 3.35 | 1.81 | 6.23 |  |  |  |  |  |
| Others work shifts | 2.83 | 1.52 | 5.32 |  |  |  |  |  |
| Asthma | 0.33 | 0.12 | 0.92 | 0.03 | 0.29 | 0.09 | 0.89 | 0.026 |
| Tobacco | 3.13 | 1.39 | 7.01 | 0.003 |  |  |  |  |
| Anxiety | 2.34 | 0.91 | 6.05 | 0.063 |  |  |  |  |

HCW: healthcare workers; h.: hours.

According to the type of mask used there was no difference in the number of days with headache in the month prior to the survey 13.4 (SD 7.4) vs 12.6 (SD 6.9), nor in the previous week 3.9 (SD 1.6) vs 3.6 (SD 1.7), but in the intensity according to VAS 5.7 (SD 1.5) vs 6.5 (1.2), p= 0.004.

The impact of headache in subjects with a filtering mask as opposed to surgery mask was worse in the four aspects evaluated by the Likert scale: occupational 4.44 vs 3.81 (p=0.206), family 5.10 vs 4.20 (p=0.065), personal 5.64 vs 4.84 (p=0.05) and social 5.46 vs 4.58 (p=0.076) (Figure 1). The impact was also greater in subjects with previous headache in the four aspects evaluated, see table 2.

**Figure 1:**
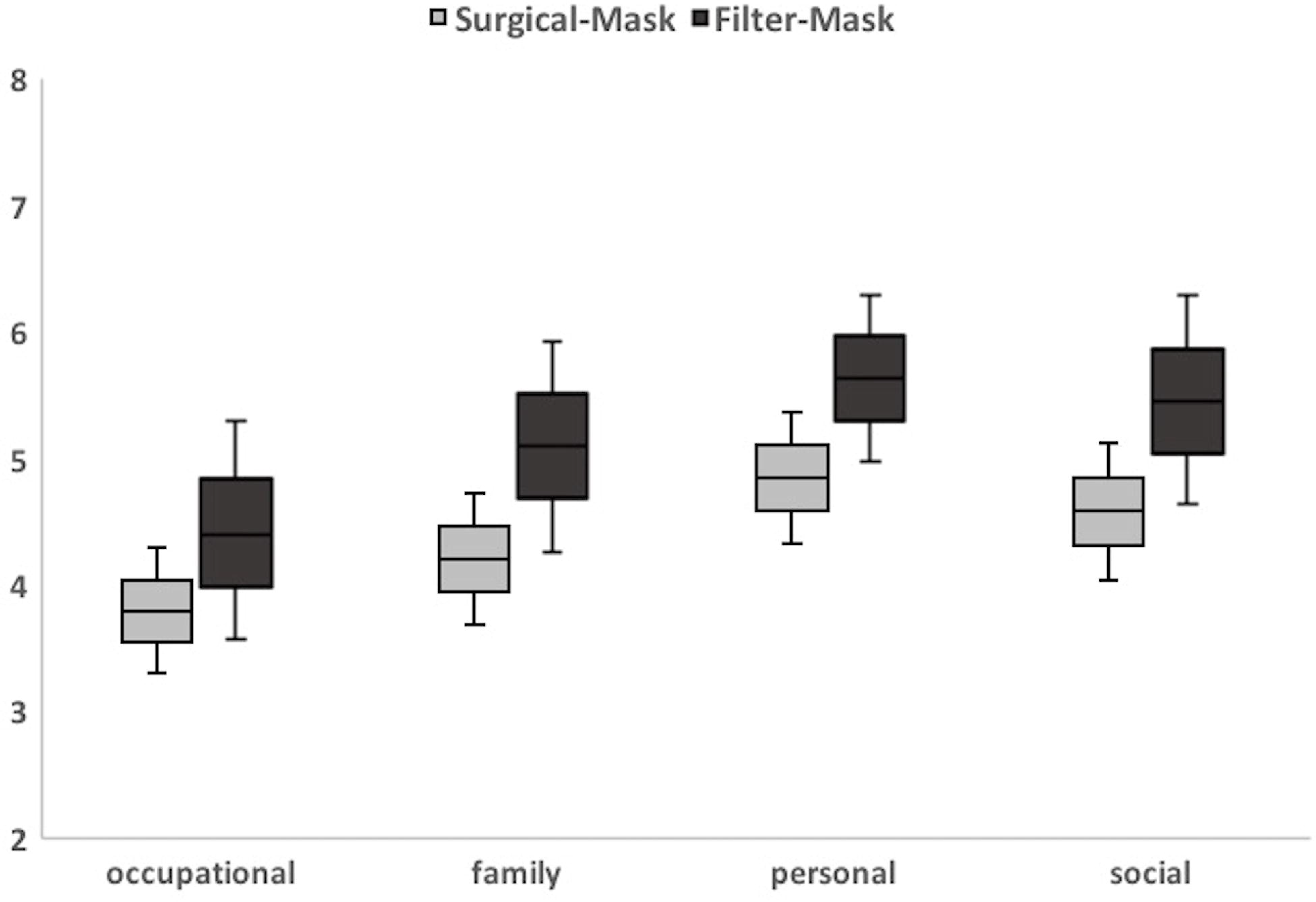
Impact of headache in subjects with a filtering mask as opposed to surgery mask in the four aspects evaluated by the Likert scale. Likert Scale Rating: Likert Scale Rating It indicates the degree of limitation due to headache in different areas of life. 0: none; 10: maximum.

Regarding the evaluation of self-perceived work stress by means of the 12 items of the PPQ, individuals with “de novo” headache versus those without headache have significantly worse scores in all aspects evaluated, except for the decrease in appetite where no significant differences are observed. Figure 2 shows graphically the evaluation of occupational stress according to the presence of headache or not. The use of a filtering mask compared with surgical mask only implies a significantly worse score in two aspects: gastrointestinal discomfort (p=0.047) and greater sensation of extreme tiredness (p=0.004).

**Figure 2:**
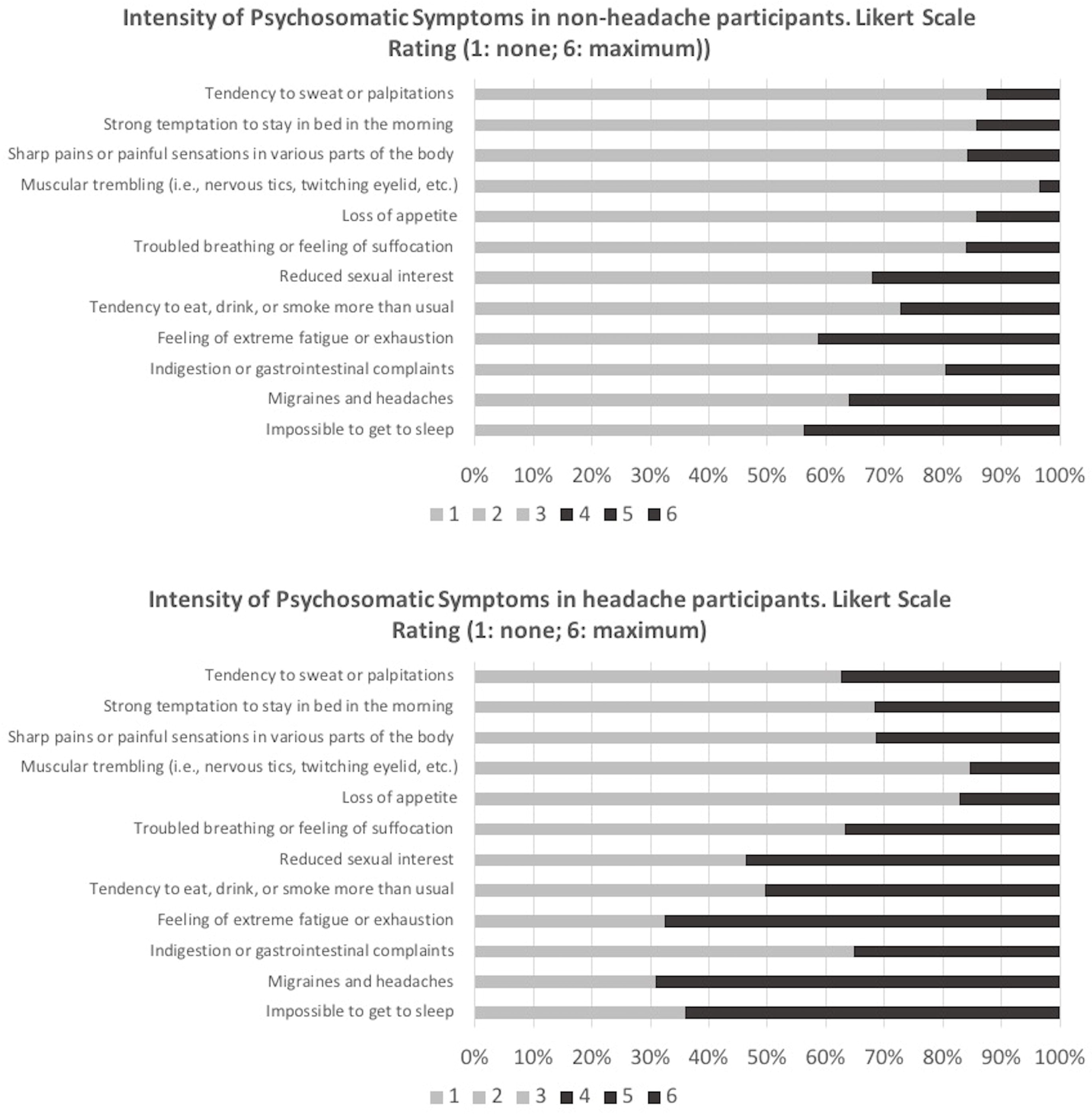
Evaluation of self-perceived work stress by means of the 12 items of the Psychosomatic Problems Questionnaire.

## Discussion

The current situation experienced by the COVID-19 pandemic has led to a substantial change in the work flows of health professionals. One of the most important features has been the use of PPE for the care of patients with suspected or infected SARS-COV2. According to the data obtained, we demonstrate a statistically significant association between the use of filtering masks and the appearance of headache.

In the physiopathology of new-onset headache, the exact mechanisms may be multiple, complex and not always well known. Peripheral nociceptive structures and central sensitization mechanisms may be involved in their development [14, 15]. The current International Headache Classification proposes, generically for secondary headaches, that the diagnostic criteria do not require remission or improvement of the underlying causal disorder before the diagnosis is formalized. There is criterion A (presence of the headache), criterion B (presence of the causal disorder) and criterion C (evidence of the etiopathogenesis). And for acute processes, a close temporal relationship between the onset of the headache and the onset of the suspected causal disorder is usually sufficient [11]. Following this classification, mask-associated headache would probably be a multifactorial disorder with unknown etiopathogenesis at present. Hypothetically, a number of factors may explain the association with filtering mask use, including hypoxia, hypercapnia, local compression and mechanical phenomena, as well as anxiety about wearing the device [10].

In the scientific literature there are not many studies that relate the use of face masks to changes in the concentration of oxygen and/or carbon dioxide but it seems a plausible hypothesis due to the barrier element that is interposed in the physiological ventilation mechanism.

In a Taiwanese cohort of 39 patients with end-stage renal disease who wore N95 masks during the 2002 SARS outbreak, they measured, among other variables, the level of Pa02 before and after a 4-hour hemodialysis session. The study concluded that there was a significant reduction in PaO2 from baseline and an increase in other respiratory adverse effects [16]. Another study conducted in a cohort of 130 astronauts subjected to high CO2 pressures during controlled training showed a significantly higher incidence of headache in the exposed group, in addition to respiratory symptoms and difficulty in concentrating [17]. At the University of Wollongong (Australia), a study on the effects of CO2 inhalation on workers wearing respiratory protection devices showed that high levels of carbon dioxide were associated with feelings of discomfort and significantly reduced tolerance and time of device use [18]. In the world of sport, the effect on respiratory physiology and muscle performance of wearing training masks designed to simulate a variable altitude situation has been studied. The results are mixed in terms of objective performance parameters, however, it does seem common that mask use reduces working speed and negatively influences levels of alertness and task focus [19]. In 2014, a pilot study evaluated the consequences on respiratory physiology of surgical mask and N-95 face mask use in a sample of 87 patients and the extent to which nasal inspiratory and expiratory resistance and discomfort were altered in the individuals. Physiological changes such as increased respiratory resistances were observed after three hours of use [20]. Headache associated with filtering mask use could be included according to ICHD-3 [11] in the section on headaches due to homeostatic disorders where those related to alteration of oxygen and carbon dioxide partial pressure parameters are included.

Another phenomenon probably related to the physiopathology of headache after PPE use is the external compression that it generates, as recently reflected by the group of Ong JJ et al [21]. In most cases there is a temporal relationship between the use of devices and the headache, as well as the topographical location of the headache. As with homeostatic changes, ICHD-3 typifies a type of headache attributable to uninterrupted compression or traction of pericranial soft tissues [11]. In this situation there is more room for the external compression subtype where the elements of the PPE (glasses or protective shields and masks, mainly N95) produce compression over several hours on different facial regions.

Kymchatowski et al. analysed a cohort of 82 military police in Rio de Janeiro exposed to the regulatory helmet, and reported headache occurrence in all cases after wearing the helmet for at least 1 hour, with 92.7% disappearing after the removal. In addition, they reported that headache was clearly different from other headaches suffered in 64.6% of the cases. One third of the sample presented migraine, referring to the fact that the new headache was more intense and completely limited the development of their activity. It was also observed in all subjects of the cohort that the headache did not reproduce after removing the stimulus for five weeks [22]. Finally, a study of 212 health professionals assessing demographic factors, time of N-95 mask use and the existence of previous headaches showed a relatively high prevalence of mask headache among health workers who worked in high-risk areas during the 2003 SARS epidemic [10].

The last factor to be mentioned is the level of anxiety or stress. Multiple ways of relating the level of stress to the occurrence of headache have been described, either as “de novo” occurrence or as exacerbation in an individual with primary headache [23]. In the case of the SARS-CoV-2 pandemic, health care workers may be affected by critical incident stress (CIS). Critical incidents are events in which people witness or experience tragedy, death, serious injury or threatening situations, which can have a strong emotional impact. The signs and symptoms of CIS can be physical, cognitive, emotional and behavioural [24]. In our work, we observed that the level of stress in headache subjects is significantly worse in all aspects measured by PPQ.

We also showed that the risk of developing headache is higher among nurses and other health professionals than among physicians. The explanation for this result is complex, but there are three plausible hypotheses. As a general rule, doctors live with a higher level of stress in the course of their work, and therefore, situations considered conflicting do not increase their usual stress threshold excessively [25]. It could also be explained by the use of negative coping strategies in some professional groups as opposed to others [26], these strategies, which we have not measured in our work, would be related to professional level. The third potential explanation, in line with some published studies, is that the higher risk of headache among nurses and other health professionals than in the medical group, is due to the differential characteristics of the workers’ occupation, which would involve the use of other devices, cleaning materials, activities with greater energy expenditure or changing work shifts [27].

Different factors or comorbidities that may influence the development of headache have been described in the literature [28]. If we look at risk markers, age and sex deserve special attention. The female sex is closely related to the development of “de novo” headache [29]. Age is a determining factor in the classification of headache according to the International Headache Society [11]. Several studies have shown that pain intensity [30], the degree of headache disability, and the possibility of secondary headache occurrence are age-related factors [31]. In terms of other individually modifiable risk factors, the relationship between blood pressure changes and primary headache should be highlighted, as they share mechanisms of action such as vascular endothelial dysfunction or poor cardiovascular autonomic regulation [32]. However, in our study we did not find a clear association between different comorbidities of the individual and the appearance of headache, except for tobacco consumption in the univariant analysis.

In a review of the relationship between smoking, its different components and the occurrence of headache, controversial data were obtained. The studies conducted in this regard are mostly retrospective and limited, and there is no definite evidence that tobacco is an independent cause of headache occurrence. However, most migraine patients define it as a trigger [33]. Headache is one of the most pronounced symptoms in patients suffering from asthma, a fact that has been described in a few studies so far. In a study of 93 patients, a statistically significant difference was found in this area, as 62.4% of asthmatics had headache (migraine or tension), whereas in the control group the percentage was only 32.8%. Other factors such as the use of steroid inhalers, the presence of rhinitis, conjunctivitis or respiratory parameters such as FEV1 were studied and characterized [34]. In our study, being asthmatic would act as a protective factor against headache associated with mask use, perhaps because of a greater tolerance to hypoxia, and therefore a higher threshold for developing headache for this reason.

## Limitations

Our study has some limitations that should be noted: the sample is one of convenience and there has been no previous probability sampling. We could not include or under-represent some professional groups. The study is cross-sectional, which helps us to formulate hypotheses, but we cannot prove causality. We have not taken into account the temporal evolution of the headache in the health professionals who present it. Nor have we taken into account other external factors that may influence the headache, such as the exact conditions of the site and type of work.

## Conclusion

In our study, we described the occurrence of “de novo” headache with the use of filtering masks and its negative impact on multiple dimensions of the life of healthcare professionals. We propose headache associated with the use of this type of mask as a new subtype of headache, of a multifactorial nature and complex etiopathogenesis. And since the use of these devices will tend to become more widespread due to the implications of the pandemic, we believe it is important to promote prevention and protection strategies that guarantee the safety of workers, without undermining their quality of life.

## Data Availability

All the data with which this work has been prepared are available to any researcher upon reasonable and understandable request to the corresponding author.

## Acknowledgements

We want to thank Juan Rodrigo Ross for his invaluable help in the preparation of this paper. Also to all colleagues, health workers who have responded with great rigor to the questionnaire.

## References

1. Zhu N, Zhang D, Wang W, Li X, Yang B, Song J, et al. A novel coronavirus from patients with pneumonia in China, 2019. N Engl J Med 2020; 382:727–33.

2. Huang C, Wang Y, Li X, et al. Clinical features of patients infected with 2019 novel coronavirus in Wuhan, China. Lancet. 2020; 395:497–506.

3. World Health Organization. Coronavirus disease (COVID-19) pandemic. Geneva: WHO. Avalaible from: https://www.who.int/emergencies/diseases/novel-coronavirus-2019 [14.05.2020]

4. National Centre for Epidemiology. Carlos III Institute of Health. Current situation COVID 19. Madrid: CNE-ISCIII. Available from: https://cnecovid.isciii.es/covid19/ [14.05.2020]

5. Ministry of Health of the Regional Government of Extremadura. Health and Social Services. Communication. “Extremadura detects 208 suspected cases of Covid-19, 3 new infections and 3 more deaths”. Available from: http://www.juntaex.es/comunicacion/noticia&idPub=30300#.Xr1-iC-b7-Y [14.05.2020].

6. Garcia-Godoy LR, Jones AE, Anderson TN, Fisher CL, Seeley KM, Beeson EA, et al. Facial protection for healthcare workers during pandemics: a scoping review. BMJ Glob Health. 2020; 5(5):e002553.

7. Van der Sande M, Teunis P, Sabel R. Professional and home-made face masks reduce exposure to respiratory infections among the general population. PLoS One. 2008; 3(7):e2618

8. Sra HK, Sandhu A, Singh M. Use of face masks in COVID-19. Indian J Pediatr. 2020; 1

9. Shenal BV, Radonovich Jr LJ, Cheng J, Hodgson M, Bender BS. Discomfort and exertion associated with prolonged wear of respiratory protection in a health care setting. J Occup Environ Hyg. 2012;9(1):59–64.

10. Lim EC, Seet RC, Lee KH, Wilder-Smith EP, Chuah BY, Ong bk. Headaches and the N95 face-mask amongst healthcare providers. Acta Neurol. Scand. 2006; 113(3):199–202

11. Headache Classification Subcommittee of the International Headache Society. The International Classification of Headache Disorders, 3rd edition. Cephalalgia 2018;38(1):1–211.

12. Sjaastad O. Headache and the influence of stress. A personal view. Ann Clin Res 1987; 19:122–8.

13. Hock RR. Professional burnout among public school teachers. Public Personnel Management. 1988 Nº 17, Vol. 2, pp. 167-189.

14. Peng KP, Fuh JL, Yuan HK, Shia BC, Wang SJ. New daily persistant headache; should migrainous features be incorporated? Cephalalgia. 2011; 31:1561–9.

15. Silberstein SD, Lipton R, Solomon S, Mathew N. Classification of daily and near daily headaches: Proposed revisions to the IHS classification. Headache. 1994; 34:1–7.

16. Kao TW, Huang KC, Huang YL, Tsai TJ, Hsieh BS, Wu MS. The physiological impact of wearing an N95 mask during hemodialysis as a precaution against SARS in patients with end-stage renal disease. J Formos Med Assoc. 2004; 103(8):624-8

17. Law J, Young M, Alexander D, Mason SS, Wear ML, Méndez CM, et al. Carbon dioxide physiological training at NASA. Aerosp Med Hum Perform. 2017; 88(10):897-902.

18. Smith CL, Whitelaw JL, Davies B. Carbon dioxide rebreathing in respiratory protective devices: influence of speech and work rate in full-face masks. Ergonomics. 2013;56(5):781–790

19. Jagim AR, Dominy TA, Camic CL, Wright G, Doberstein S, Jones MT, et al. Acute effects of the elevation training mask on strength performance in recreational weight lifters. J Strength Cond Res. 2018; 32(2):482-489

20. Zhu JH, Lee SJ, Wang DY, et al. Effects of long-duration wearing of N95 respirator and surgical facemask: a pilot study. J Lung Pulm Respir Res. 2014;1(4):97–100.

21. Ong JJ, Bharatendu C, Goh Y, Tang JZ, Sooi KW, Tan YL, et al. Headaches associated with personal protective equipment – A cross-sectional study among frontline healthcare workers during COVID-19. Headache. 2020; 60(5):864-877

22. Krymchantowski AV. Headaches due to external compression. Curr Pain Headache Rep. 2010; 14:321–324

23. Nash JM, Thebarge RW. Understanding psychological stress, its biological processes, and impact on primary headache. Headache. 2006; 46 (9); 1377-1386.

24. Lowe SR, Bonumwezi JL, Valdespino-Hayden Z, Galea S. Posttraumatic Stress and Depression in the Aftermath of Environmental Disasters: A Review of Quantitative Studies Published in 2018. Curr Environ Health Rep. 2019;6(4):344–360.

25. Houle T, Nash JM. Stress and Headache Chronification. Headache. 2008; 48(1); 40–44.

26. Xu HG, Johnston ANB, Greenslade JH, Wallis M, Elder E, Abraham L, et al. Stressors and coping strategies of nurses and emergency department doctors: a cross-sectional study. Australas Emerg Care. 2019; 22(3):180-18

27. Pucci R, Matozzo F, Arrigo A, Mazza S, Sandrini G, Nappi G. Prevalence of primary headache realted to work activity in a group of hospital workers undergoing periodic visits. G Ital Med Lav Ergon. 2003; 25(4); 448-452.

28. Jensen R, Stovner LJ. Epidemiology and comorbidity of headache. Lancet Neurol. 2008; 7(4); 354–361.

29. Stovner LJ, Andree C. Prevalence of headache in Europe: a review for the Eurolight project. J Headache Pain. 2010; 11(4); 289–299.

30. Song TJ, Kim YJ, Kim BK, Kim BS, Kim JM, Kim SK, et al. Characteristics of Elderly-Onset (≥65 years) Headache Diagnosed Using the International Classification of Headache Disorders, Third Edition Beta Version. J Clin Neurol. 2016; 12(4); 419-425

31. Guido D, Leonardi M, Mellor-Marsá B, Moneta MV, Sanchez-Niubo A, Tyrobolas S, et al. Pain rates in general population for the period 1991–2015 and 10-years prediction: results from a multi-continent age-period-cohort analysis. J Headache Pain. 2020; 21(1):52

32. Guan T, Hu S, Han Y, Wang R, Zhu Q, Hu Y, Fan H, Zhu T. The effects of facemasks on airway inflammation and endothelial dysfunction in healthy young adults: a double-blind, randomized, controlled crossover study. Part Fibre Toxicol. 2018 Jul 4;15(1):30. doi: 10.1186/s12989-018-0266-0.

33. Taylor FR. Tobacco, Nicotine, and Headache. Headache. 2015;55(7):1028–1044.

34. Gungen AC, Gungen B. Assessment of Headache in Asthma Patients. Pak J Med Sci. 2017;33(1):156–161.

